# Unrestricted Online Sharing of High-frequency, High-resolution Data on SARS-CoV-2 in Wastewater to Inform the COVID-19 Public Health Response in Greater Tempe, Arizona

**DOI:** 10.1101/2021.07.29.21261338

**Authors:** Devin A. Bowes, Erin M. Driver, Simona Kraberger, Rafaela S. Fontenele, LaRinda A. Holland, Jillian Wright, Bridger Johnston, Sonja Savic, Melanie Engstrom Newell, Sangeet Adhikari, Rahul Kumar, Hanah Goetz, Allison Binsfeld, Kaxandra Nessi, Payton Watkins, Akhil Mahant, Jake Zevitz, Stephanie Deitrick, Philip Brown, Richard Dalton, Chris Garcia, Rosa Inchausti, Wydale Holmes, Xiao-Jun Tian, Arvind Varsani, Efrem S. Lim, Matthew Scotch, Rolf U. Halden

**Author notes:** These authors contributed equally (Devin A. Bowes, Erin M. Driver).

## Abstract

The COVID-19 pandemic prompted a global integration of wastewater-based epidemiology (WBE) into public health surveillance. Among early pre-COVID practitioners was Greater Tempe (population ~200,000), Arizona, where high-frequency, high-resolution monitoring of opioids began in 2018, leading to unrestricted online data release. Leveraging an existing, neighborhood-level monitoring network, wastewater from eleven contiguous catchment areas was analyzed by RT-qPCR for the SARS-CoV-2 E gene from April 2020 to March 2021 (*n*=1,556). Wastewater data identified an infection hotspot in a predominantly Hispanic and Native American community, triggering targeted interventions. During the first SARS-CoV-2 wave (June 2020), spikes in virus levels preceded an increase in clinical cases by 8.5±2.1 days, providing an early-warning capability that later transitioned into a lagging indicator (−2.0±1.4 days) during the December/January 2020-21 wave of clinical cases. Globally representing the first demonstration of immediate, unrestricted WBE data sharing and featuring long-term, innovative, high-frequency, high-resolution sub-catchment monitoring, this successful case study encourages further applications of WBE to inform public health interventions.

---

Triggered by the SARS-CoV-2 pandemic, the use of wastewater-based epidemiology (WBE) as a potentially powerful, rapid, and inexpensive tool to inform public health decision-making has seen a remarkable increase globally. For decades, WBE has been exercised to track chemical and biological threats, with numerous studies underscoring its efficacy and usefulness for understanding and managing community health ^1–8^. At the onset of the SARS-CoV-2 pandemic, significant delays in conventional and individualized clinical testing, due in part to an overwhelmed healthcare system and resource limitations ^9^, positioned WBE as a promising supplemental tool for assessing SARS-CoV-2 spread at the population-level, a strategy that soon was adopted more broadly ^10–13^. Early data showed SARS-CoV-2 levels in wastewater and sludge as a concomitant or early indicator of clinical confirmed infections, disease and mortality in a community ^14, 15^.

The City of Tempe, Arizona, residential population ~200,000, had been an early adopter of WBE for the purpose of tracking opioid consumption, which began in May of 2018 and led to the launch of a fully interactive, public-facing, open access WBE dashboard in February of 2019 ^16^. In a municipal-academic partnership, Tempe and Arizona State University (ASU) participated in sharing of monthly wastewater samples, subsequent analysis, and to joint reporting of use-trends of opioids within the community monthly by displaying the obtained collaborative results for oxycodone, codeine, heroin, and fentanyl (and metabolites; µg d^-1^ per 1000 people) in five urban sewersheds ^17^. The City also had established a routine for data analysis and public health response by integrating Tempe Fire Medical Rescue, Human Services (e.g., CARE 7 crisis intervention organization), and others into a workgroup that relied on WBE data as an important and innovative source of information to guide resource deployment by community need (**Figure 1**).

With this existing framework in place, Tempe and ASU were in a unique position at the start of the SARS-CoV-2 pandemic to quickly transition into WBE surveillance of SARS-CoV-2. Coincidentally, Tempe and the ASU community also had one of the first early diagnoses of a positive SARS-CoV-2 patient (26 January 2020)^18^. As the City of Tempe and ASU quickly transitioned into molecular-based monitoring, the immediate goal was to use previously established expertise in sampling, infrastructure access, and WBE-framed public health response to begin quantitative assessments of SARS-CoV-2 levels in wastewater. The ultimate objective was to identify hotspots of infection early and implement interventions including education, outreach, and targeted clinical testing to limit the spread of the virus within the Greater Tempe community. The local health department shared data from clinical testing of individuals only at the zip code level, ^19^ a policy intended to protect small communities and personal identifiable information, which potentially limited stakeholders’ ability to respond to local virus clusters. Unique to the US, zip codes are a series of 5 numbers created by the US postal service to delineate small geographical areas within counties to improve mail service, and are used extensively by local and state agencies, including public health departments ^20^. The 5-digit, Tempe, AZ zip codes involved in this study are 85281, 85282, 85283, and 85284, and will be referred to here as ZC-1, ZC-2, ZC-3, and ZC-4. The pre-existing, neighborhood-level wastewater monitoring network offered an opportunity to test the potential of WBE to serve as an early warning system that may reveal virus presence and spread prior to clinical case data reported from testing of individuals ^21,22^. Thus, important goals of the work were (i) to compare WBE data to newly reported clinical cases of SARS-CoV-2, related hospitalizations, and associated deaths at a high temporal and geospatial resolution (i.e., county, city, zip code, and neighborhood levels), and (ii) to determine whether the concurrent pandemic monitoring by WBE produced data and information not available or obvious from clinical testing.

**Figure 1.**
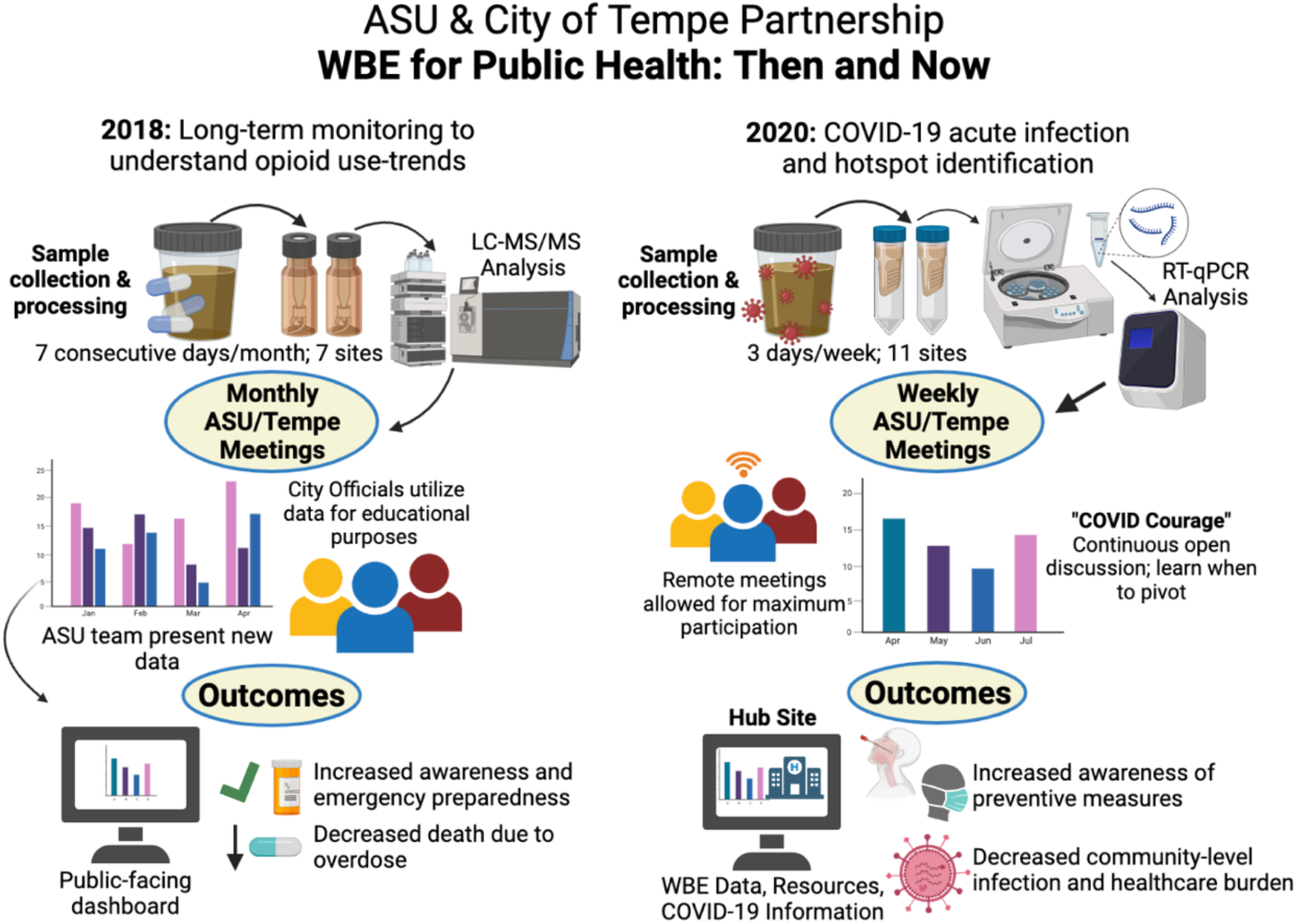
Schematic of the ASU/Tempe partnership, demonstrating how the existing wastewater monitoring network for opioid use (established in 2018) enabled a rapid transition to monitoring SARS-CoV-2 during the COVID-19 global pandemic (2020) with work products including the world’s first WBE-informed public interactive online dashboards to combat the opioid and COVID-19 epidemics through a data-driven targeted public health response. Created with BioRender.com.

## Results

### Neighborhood-level Sampling

At the onset of the pandemic, our team had divided the Greater Tempe area into five sewer catchments (Areas 1-5; **Figure 2a**), including two additional, non-published locations that received wastewater from adjacent municipalities, which were necessary to determine the Tempe-associated sewage signal where wastewater was comingled. The neighborhood-level sampling methodology was synchronized with reoccurring compliance monitoring of the Sub-Regional Operating Group (SROG), a cohort of five municipalities including Phoenix, Tempe, Mesa, Glendale, and Scottsdale, that jointly own and operate the 91^st^ Avenue wastewater treatment plant (WWTP) in Phoenix, Arizona. The predefined sampling strategy consisted of 7-consecutive days of sample collection each month, across variable weeks from permanent, sub-surface sampling stations. While this sampling strategy was sufficient for long-term, opioid-related monitoring, tracking of SARS-CoV-2 levels required an increased temporal resolution. Accordingly, we adopted a high-frequency sampling approach consisting of weekly collection on Tuesday, Thursday, and Saturday, in addition to the SROG sampling events. To improve spatial resolution, additional sampling locations were also identified based on ease of collection (Area 6), while three other permanent locations needed slight infrastructure modifications (Area 7) and/or approvals prior to onboarding, including the Town of Guadalupe with strong representation by Native American and Hispanic residents (**Figure 2a**), and Tempe St. Luke’s Hospital (not displayed on dashboard). Permanent sampling locations outside of the Tempe jurisdiction (necessary for eliminating non-Tempe SARS-CoV-2 signals) were available only during the previously referenced week of compliance monitoring. City of Tempe personnel began sampling from maintenance holes (also known as manholes) immediately downstream of these locations, within the City’s jurisdiction, during the three weeks each month when regular compliance sampling was not performed.

**Figure 2.**
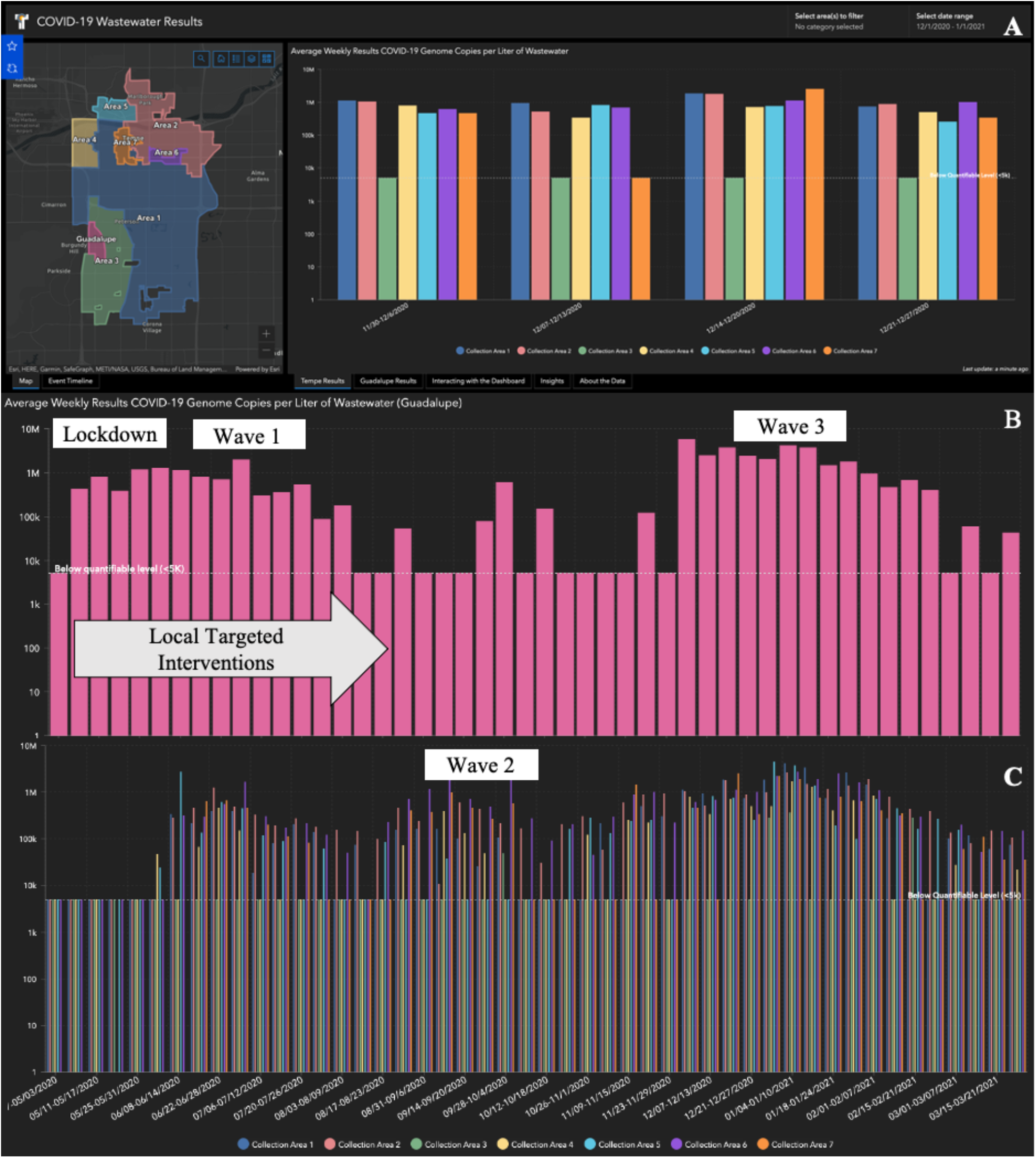
Publicly accessible, interactive WBE online dashboard showcasing (A) a Map of Tempe, Arizona divided into wastewater catchments (Areas 1-7 and Town of Guadalupe) alongside one month of SARS-CoV-2 levels in wastewater determined by targeting the E gene, with additional data being displayed for (B) The Town of Guadalupe (May 2020 through March 2020); with local targeted interventions implemented from May 2020 through August 2020 and (C) The City of Tempe (May 2020 through March 2020), showing the impact of the initial lockdown, and subsequent waves of infection over time (viewable at covid19.tempe.gov).

### Determination of SARS-CoV-2 Levels in Wastewater

Between 1 April 2020 and 31 March 2021, a total of 1,556 samples were collected across the Greater Tempe area. Each 24-h composite sample collected represented either a flow-weighted or time-averaged sample (15 min. collection intervals) captured by a high-frequency automated sampler. The number of samples collected per catchment during the study varied from a high of 155 in Area 1 to lows of 103 (Area 7) and 101 (Hospital), with observed differences resulting from occasional sampler malfunctioning and staggered onboarding of additional sampling locations. The total number of SARS-CoV-2 detects per catchment throughout the study was on average 66 ± 36, with a minimum of 4 (Area 3) to a maximum of 116 (Area 6). On average, when detected, the coronavirus concentration was 617,000 ± 2.075 million (M) gene copies L^-1^ (median of 251,450 gene copies L^-1^) indicative of great fluctuations in virus levels over time. Detailed concentration information is provided in **Figure S1**.

The SARS-CoV-2 viral load was calculated for each sample at a collection point using wastewater flow data provided by Tempe (**Figure S2**). Flow rates in catchment Areas 1-7 had data recorded at 2-min intervals in real-time using permanent laser flow meters, while the Town of Guadalupe and the Tempe St. Luke’s Hospital had only historical flow data available. Flow varied from a maximum in Area 1 of 54.5 ± 6.6 million L day^-1^ (MLD) to a minimum of 0.106 MLD (historical estimate) for the hospital location; average flow rates across all catchments were 15.1 ± 21.3 MLD.

At select collection sites, the corresponding wastewater sample was representative of multiple collection catchment areas due to the comingling of wastewater in the collection system (**Figure S3**); this occurred in Areas 1-3. To isolate individual catchments and provide a catchment-specific viral load, a mass balance was performed. Resultant viral loads in each sewer catchment ranged from 6 × 10^10^ - 1 × 10^13^ genome copies d^-1^ (**Figure S4**). The distributions in viral load varied between each location, with Areas 1, 2, and 6 showing smoother distributions in viral load over time, while others showed more isolated single-day spikes in activity. Most locations showed two waves in viral levels occurring in June 2020 and December/January 20-2021. However, in catchments close to ASU, an additional unique wave of viral load was visible (Areas 6 and 7) in August 2020 (**Figure S4**).

Understanding the number of people contributing to wastewater in any given catchment is critical when working with data generated from within the collection system. Estimated Tempe subpopulations ranged from a low of 8,114 ± 848 in Area 5 to a high of 132,082 ± 7,374 in Area 1, the largest geographic catchment area (**Table S1**). Variability in Tempe data was a function of the total numbers of residents, employed individuals, and the number of students in the contributing area. The population of the Town of Guadalupe (6,500) was determined using US census data ^23^. The hospital location was omitted from this population analysis since the number of individuals working or serving as patients was unknown.

### Data Usage for Public Health

The result of these efforts ultimately culminated in eight SARS-CoV-2 collection locations viewable online by the public (Areas 1-7 of Tempe and Town of Guadalupe) on an interactive dashboard that went live the first week of May 2020 (**Figure 2**). The dashboard displays each catchment area overlain on a street-level city map so users can geospatially identify contributing locations in the catchment. In response to a request of the impacted communities, the Town of Guadalupe is displayed on a separate tab of the dashboard. Data are shown as the logarithm of genome copies L^-1^ and are presented as a weekly average consisting of the Tuesday, Thursday, and Saturday collected samples. Since the sewage collection system in Tempe separates stormwater from municipal wastewater and the study site is in an arid climate, the use of concentration was permissible. Users have the ability to control spatial and temporal parameters to their preference; and text and infographics accompany these data, which explain WBE basics, how to properly interpret the data, and how data are created and used by the City. Additionally, the SARS-CoV-2 wastewater dashboard is nested in a Community COVID-19 Health Site that contains information provided by the Centers for Disease Control and Prevention (symptoms, prevention, exposure-response), City demographic information, and positive clinical cases reported by zip code. To ensure congruency in data interpretation between Tempe and ASU, the frequency of joint meetings that began in 2018 increased from monthly to weekly beginning April, 2020 or biweekly (December, 2020 and on) to discuss wastewater-derived data, newly reported cases in the community, sampling logistics, and targeted mitigation strategies in communities in response to the collected data when applicable. Although sewage temperature, travel time, and storage are known to influence the stability and signal strength of labile wastewater-borne biomarkers such as virus RNA ^24^, a lack of temperature-specific attenuation rates of SARS-CoV-2 did not allow for a confident normalization of data during the time of study. However, the short residence times within the various neighborhood-level sewersheds in Tempe and use of refrigerated samplers decreased the impact of these variables on collected samples.

Data generated from the SARS-CoV-2 wastewater monitoring revealed consistently elevated virus measurements in the Town of Guadalupe during the initial lockdown in May to early June 2020, a marked difference compared to the other catchments in the area. A targeted approach of implementing face-mask mandates and community education in town halls, adding clinical testing sites, in addition to public sharing of wastewater data occurred from April through August, 2020, resulting in the decline of both viral loading in wastewater and newly reported SARS-CoV-2 positive cases. The Town of Guadalupe’s wastewater comingles with Tempe wastewater (and an additional external municipality) and is collected as a composite at Area 3. However, Area 3 only had four detects during the entire year-long sampling campaign, implying that the elevated SARS-CoV-2 signal originating in the Town of Guadalupe was attenuated beyond detectability at the Area 3 sampling location, and was visible only through high-resolution monitoring. Whereas clinical testing in theory may have rendered the Guadalupe infection hotspot visible, testing was lacking in the area, and newly reported case counts from traditional clinical tracking by the county were aggregated at the zip code level with that of Tempe, thereby obscuring what may have been happening within the community.

## Discussion

We employed WBE to monitor SARS-CoV-2 in the Greater Tempe, Arizona, Southwestern United States by implementing a unique, high-frequency, and neighborhood-level sampling approach in conjunction with immediate, open access data sharing with the public. The present work illustrates how an established WBE monitoring network can be adopted quickly to shift from one public health priority to another, as done here by switching from opioid targets to SARS-CoV-Measured values in this study were in line with those reported from other wastewater monitoring studies; ^15, 25, 26^ however, the maximum concentration of 37.6 M gene copies L^-1^, is among the highest recorded measured values to date. This measurement occurred at the hospital location, which had an active COVID-19 ward at the time of collection on 11 January 2021 during peak pandemic conditions (to date). Higher relative standard deviations (RSD) in measured SARS-CoV-2 values for a given week (*n*=3 observations per week), occurred in locations with a higher proportion of commercial businesses, including Areas 4 and 5 (RSD 83, 93%) as compared to those with largely residential catchments (Areas 1 [58%] and Area 2 [65%]). This may explain the relatively smoother trends over time in Areas 1 and 2, as compared to those with higher transient populations, which showed isolated single-day spikes in viral presence. This is plausible given non-ambulatory individuals likely would have stayed at home throughout the duration of their illness. These results suggest that a high-frequency sample collection approach should be considered in catchments with a higher proportion of transient populations, which may be susceptible to greater variability in virus occurrence from day to day.

Estimating population size by study area was challenging due to the unique nature of collecting wastewater from within the sewer infrastructure rather than by determining the population by counting the residents of local buildings served by a wastewater treatment plant as performed in traditional WBE studies ^27, 28^. As a net importer of people to the City for work, it was important not only to quantify the residents but also the non-resident employed and transient student populations, a task accomplished by using Maricopa Association of Governments (MAG) and on-campus student resident data provided by ASU. MAG data needed to be corrected for lockdown activities which closed businesses, for which we used Arizona department of transportation arterial traffic flow data (e.g., 40% decrease in arterial traffic equated to 40% decreased in employment populations). Due to the bulk of student classes moving online, using only campus resident data (based on student housing contracts which were updated monthly) was appropriate to assess temporal changes in student populations. These numbers did not account for changes in resident population during holiday travel or off-campus housing locations; thus, overall percentage changes in wastewater flow were also used to estimate population size changes. For instance, wastewater flow from Area 6 increased by 20% and was sustained throughout the academic year; therefore, the population in that area was assumed to increase proportionally. This increase in total flow in Area 6 also coincided with increases in viral load, suggesting that infected students were moving back into Tempe for the start of the academic year. Looking ahead to Fall 2021 when classes are expected to resume in-person, quantifying that transient population will become more important for sewersheds impacted by students. We have therefore begun testing the utility of campus Wi-Fi data to better estimate population size as students and faculty return to pre-pandemic campus activities ^10^.

The measured viral loads per day of SARS-CoV-2 within each catchment area in Tempe were aggregated and partitioned to their respective zip codes (ZC-1 through 4) according to their estimated percent contribution (**Figure S5)**. Wastewater-derived SARS-CoV-2 peaks in activity correlated with newly detected clinical cases per day in three distinct waves of activity: June 2020, August 2020, and December/January 2020-21. ZC-1, home to ASU, was the only zip code that showed viral increases in August 2020. Contributions to viral load within a given community by university students, however, is not an event isolated to Tempe ^29–31^. Comparisons between spikes of coronavirus levels in wastewater and clinical case data showed that peaks in wastewater preceded positive clinical cases by 7, 6, 11, and 10 days for ZC-1 through 4 (average of 8.5 ± 2.1 days), during the first wave of the pandemic, and again during the isolated university-associated wave, this time by 6 days (**Figure 3**). These results align with preliminary assessments of wastewater and clinical case data that suggested monitoring wastewater provided an early-warning capacity ranging between 2 and 21 days ^15, 25, 32^. Tempe aggregated viral loads were also compared to Maricopa County Public Health data (**Figure 4**). Results again showed peaks in wastewater measurements preceded new clinically reported cases, SARS-CoV-2-related hospitalizations, and SARS-CoV-2-related deaths by 2, 16, and 18 days during the first wave of the pandemic. These results align with prior work demonstrating that wastewater can serve as an early indicator of future clinical case load, morbidity, and mortality.

**Figure 3.**
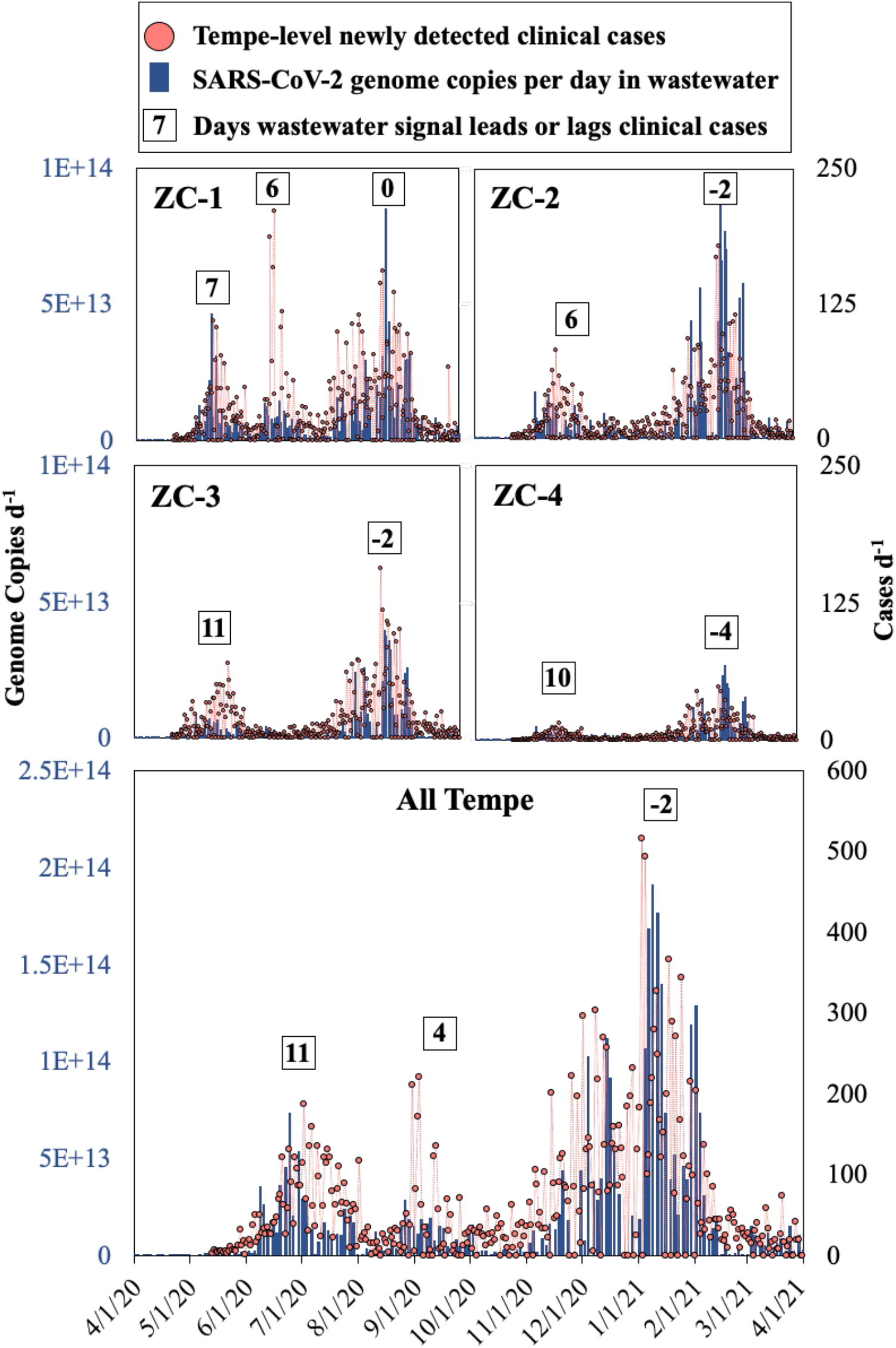
SARS-CoV-2 genome copies per day in the four zip codes (ZC-1 through 4) of Tempe, AZ and in aggregate, overlaid with newly reported clinical SARS-CoV-2 cases. Boxed numbers are the number of days the wastewater signal leads (+) or lags (-) clinical cases, determined by root mean square error (RMSE) analysis.

**Figure 4.**
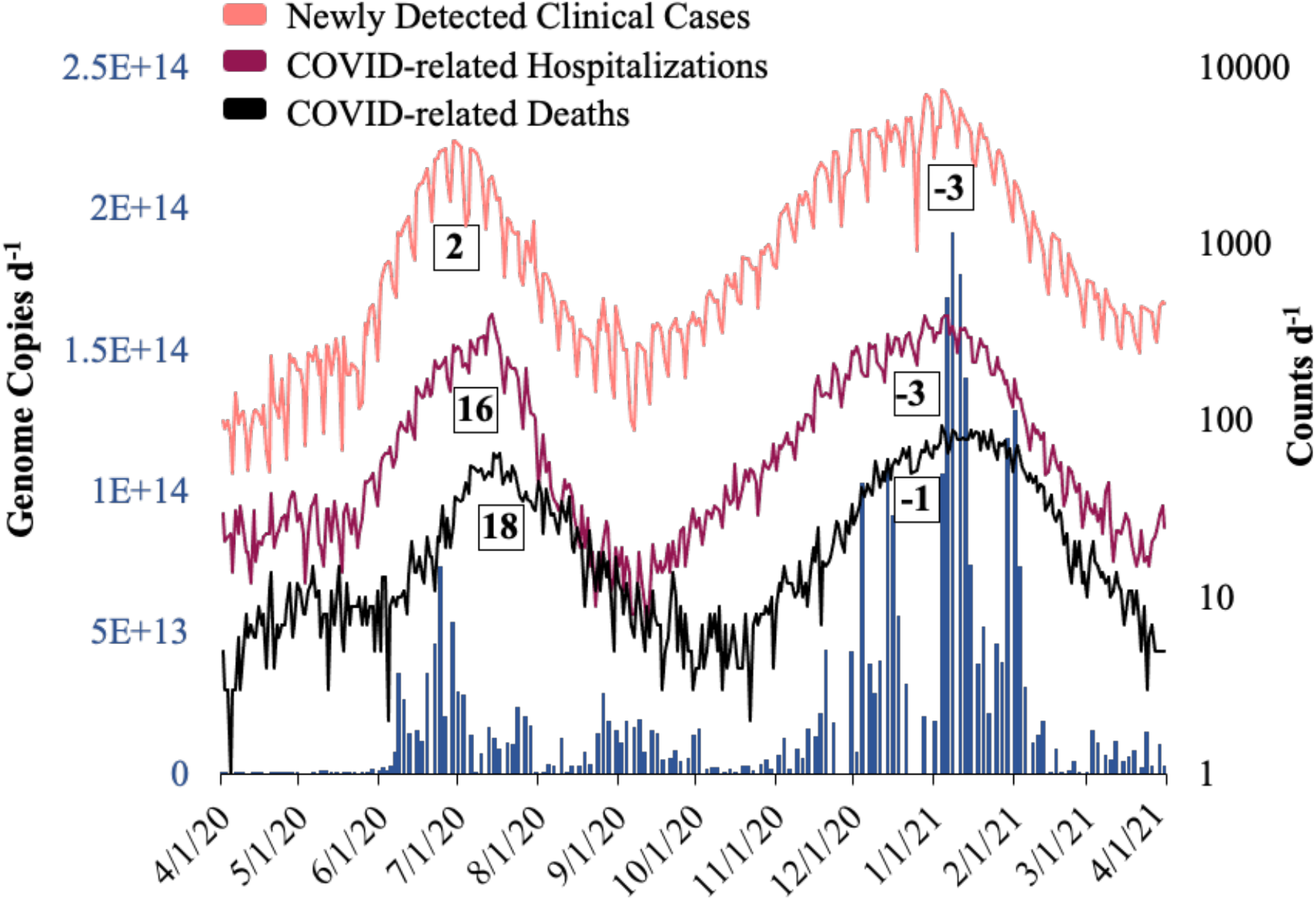
Peaks in SARS-CoV-2 viral load (genome copies d^-1^) in Tempe, AZ wastewater as compared to Maricopa County, AZ new clinically detected cases, SARS-CoV-2-related hospitalizations, and related deaths. Boxed numbers display days the wastewater signal leads (+) or lags (-) clinical cases.

Interestingly, four months later during the December/January 2020-21 wave of the pandemic, wastewater was no longer a leading indicator in any region in Tempe, AZ. Trends either directly aligned with newly reported clinical cases (ZC-1) or lagged behind clinical case data by 2 days (ZC-2 & 3), 4 days (ZC-4), and 2 days (aggregate). At the county level, wastewater lagged behind clinical results with wastewater peaking 3, 3, and 1 days behind Maricopa reported cases, SARS-CoV-2-related hospitalizations, and deaths, respectively. To our knowledge, no study has reported wastewater as both a leading and a lagging indicator to clinical cases or demonstrated such a transition from lead to lag at the sub-catchment level within the same community and from within the sewer collection system. This consecutive decrease in lead time between wastewater measurements and clinical testing may best be explained by notable differences in the availability and frequency of clinical testing over the course of this study. Qualitative data from Maricopa County shows that access to testing was extremely limited during the early stages of the pandemic ^33, 34^ but increased dramatically subsequently with the continuous onboarding of commercial, hospital, and university laboratories, largely driven by ASU biomedical screening. Thus, these data strongly suggest that the greatest benefits of WBE are to be verified early on during the detection of disease outbreaks before health care providers can mount a response. Similar benefits may be reaped late into an epidemic, when clinical testing of individuals becomes cost-prohibitive and may appear unproductive when generating mostly negative results.

This long-term study constitutes a powerful demonstration of employing WBE to collect open access, actionable data that were shown here to directly help inform and shape the public health response. High-throughput monitoring of the E gene of SARS-CoV-2 in Tempe sewage showed WBE to provide an early-warning benefit, particularly in smaller subpopulations, with a temporal and spatial data resolution that exceeded that of clinical healthcare data, which are shared only to a limited degree with local stakeholders. Use of WBE may also be important for communities with barriers to testing (e.g., lack of access, deficit of testing locations, cost), and testing fear (disbelief), or apathy as the duration of the pandemic continues and vaccination levels rise. Most importantly, WBE performed with a high spatial resolution was demonstrated to increase the ability to identify and localize hotspots of infection, thereby allowing for resources and interventions to be implemented in a targeted and more productive fashion.

Sharing data of significant economic and public health importance in a real time, open access format is often considered controversial, potentially leading to apprehension. However, leading up to, and during the study, the City’s commitment to open communication in town halls and open-attendance meetings increased transparency and trust from the community. The actions and public health outcomes achieved with this strategy here certainly appear to have outweighed any potential concerns. As such, the City of Tempe is now exploring the applicability of this methodology to other general markers of community behavior and health status. The lessons reported here may inform other communities interested in adopting this new approach, serving as a foundational framework for integrating WBE into public health monitoring and the design and implementation of intervention strategies.

## Data Availability

The SARS-CoV-2 Wastewater Monitoring Dashboard and COVID-19 Community Health Site in the City of Tempe, Arizona is available for public use at the link provided.

https://www.covid19.tempe.gov

## General

We thank the City of Tempe for their diligent collection of wastewater samples for this project throughout the course of the COVID-19 pandemic. We also thank ASU students N. Biyani and I. Mondal for their help in sample pickups, as well as E. Clancy, A. Yanez, C. Rogers, B. McFayden, M. Shope for their help with sample processing.

## Author contributions

DAB, EMD: conceptualization, investigation, sample processing, data generation and analysis, writing - original manuscript draft

SK, RF, LAH, AV, ESL: method development, sample processing and analysis, manuscript review

HG: data analysis

JW, BJ, SS, MN, SA, RK, AB, KN, PW, AM, JZ: sample processing

SD, PB, RD, CG: external data generation

RI, WH: Tempe program management

XT: data analysis oversight, manuscript edits AV, ESL, MS: oversight, manuscript edits

RUH: study conception, proposal, implementation, and data reporting

## Data and materials availability

SARS-CoV-2 Wastewater Monitoring Dashboard and COVID-19 Community Health Site in the City of Tempe, AZ: covid19.tempe.gov

