## Supplementary Information for "Unrestricted Online Sharing of High-frequency, High-resolution Data on SARS-CoV-2 in Wastewater to Inform the COVID-19 Public Health Response in Greater Tempe, Arizona"

**Supplementary Materials**

**Table S1**. Population estimates for the seven catchments in Tempe and Guadalupe. Tempe estimates were based on 2010 census data, employment data from the Maricopa Association of Governments, and Arizona State University student population. Note that Tempe St. Luke's Hospital was not included here as patient and provider information were not provided.

| Year | Month | Area 1 | Area 2 | Area 3 | Area 4 | Area 5 | Area 6 | Area 7 | Guadalupe |
|---|---|---|---|---|---|---|---|---|---|
| **2020** | April | 117,769 | 47,882 | 42,124 | 10,498 | 6,322 | 7,449 | 9,382 | 6,500 |
| | May | 127,137 | 53,788 | 47,333 | 13,582 | 7,736 | 7,588 | 10,763 | 6,500 |
| | June | 129,578 | 55,327 | 48,690 | 14,386 | 8,104 | 7,624 | 11,122 | 6,500 |
| | July | 127,662 | 54,119 | 47,625 | 13,755 | 7,815 | 7,595 | 10,840 | 6,500 |
| | August | 133,533 | 58,837 | 48,033 | 13,997 | 7,926 | 9,356 | 12,372 | 6,500 |
| | September | 137,354 | 61,395 | 50,076 | 15,206 | 8,480 | 9,423 | 12,984 | 6,500 |
| | October | 138,664 | 62,272 | 50,776 | 15,621 | 8,670 | 9,446 | 13,193 | 6,500 |
| | November | 139,537 | 62,857 | 51,243 | 15,898 | 8,797 | 9,461 | 13,333 | 6,500 |
| | December | 133,855 | 63,406 | 51,068 | 15,794 | 8,749 | 8,686 | 11,753 | 6,500 |
| **2021** | January | 138,060 | 59,551 | 51,170 | 15,854 | 8,777 | 9,766 | 11,898 | 6,500 |
| | February | 141,574 | 61,817 | 53,067 | 16,978 | 9,292 | 9,830 | 12,406 | 6,500 |
| | March | 142,520 | 64,521 | 53,578 | 17,280 | 9,430 | 9,848 | 12,542 | 6,500 |

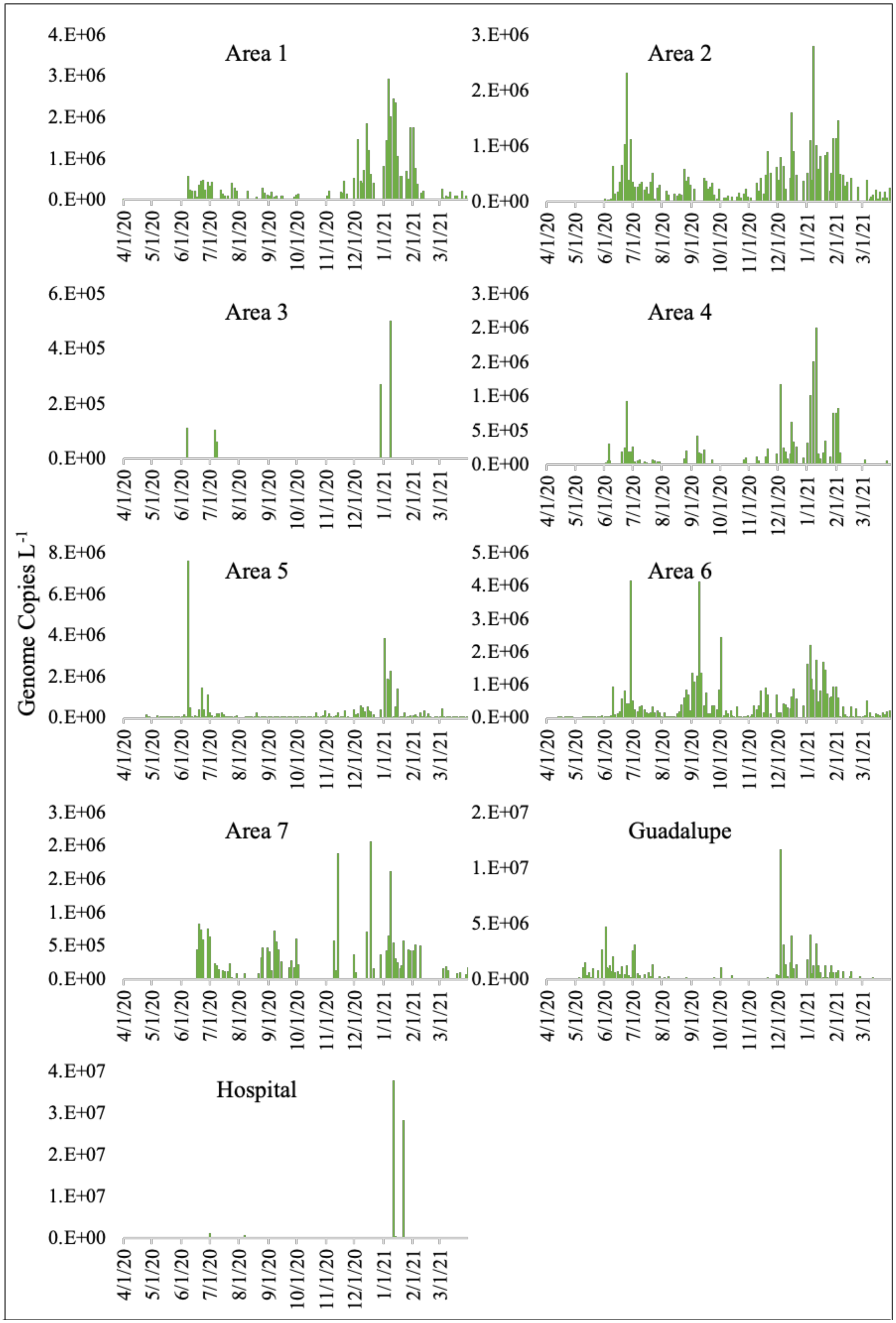


**Figure S1**. Measured concentration in target catchments from 1 April 2020 – 31 March 2021.

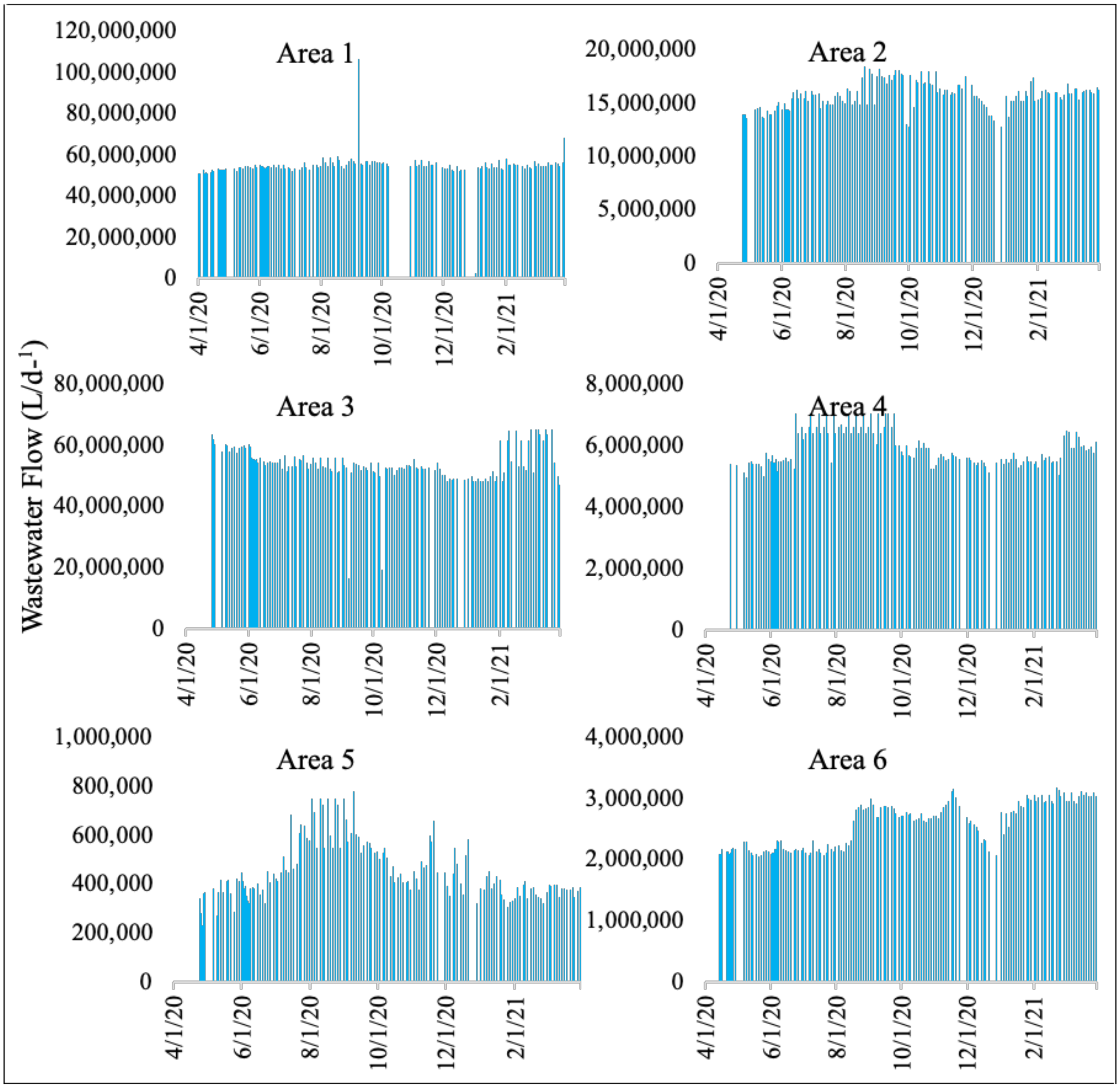


**Figure S2.** Daily flow wastewater measurements for Tempe catchments. Data are only shown for days when samples were collected. Guadalupe and the Tempe St. Luke's hospital location used historical flow data, 370,000 L $d^{-1}$ and 105,992 L $d^{-1}$ respectively and are not shown here.

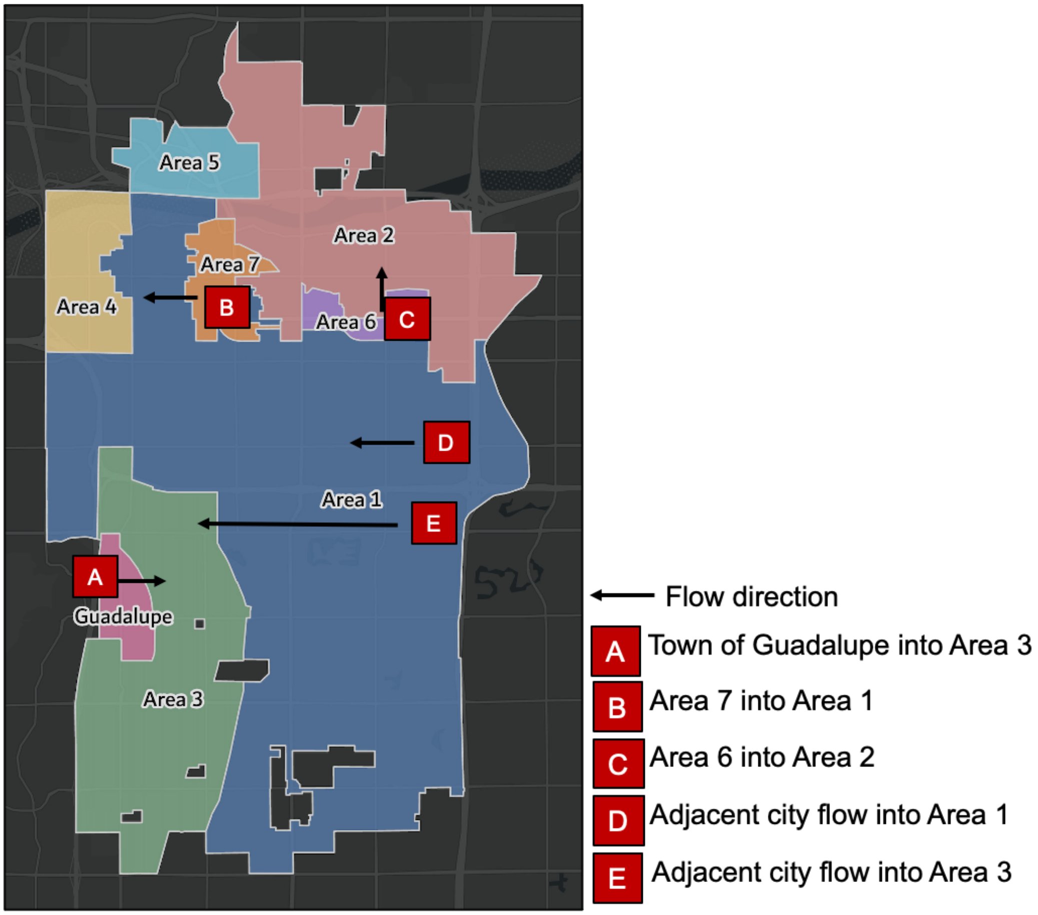


**Figure S3.** Wastewater flow direction in target catchment areas that illustrate the comingling of waters either from other Tempe-specific catchments or adjacent city flow. Relevant locations include Areas 1, 2, and 3.

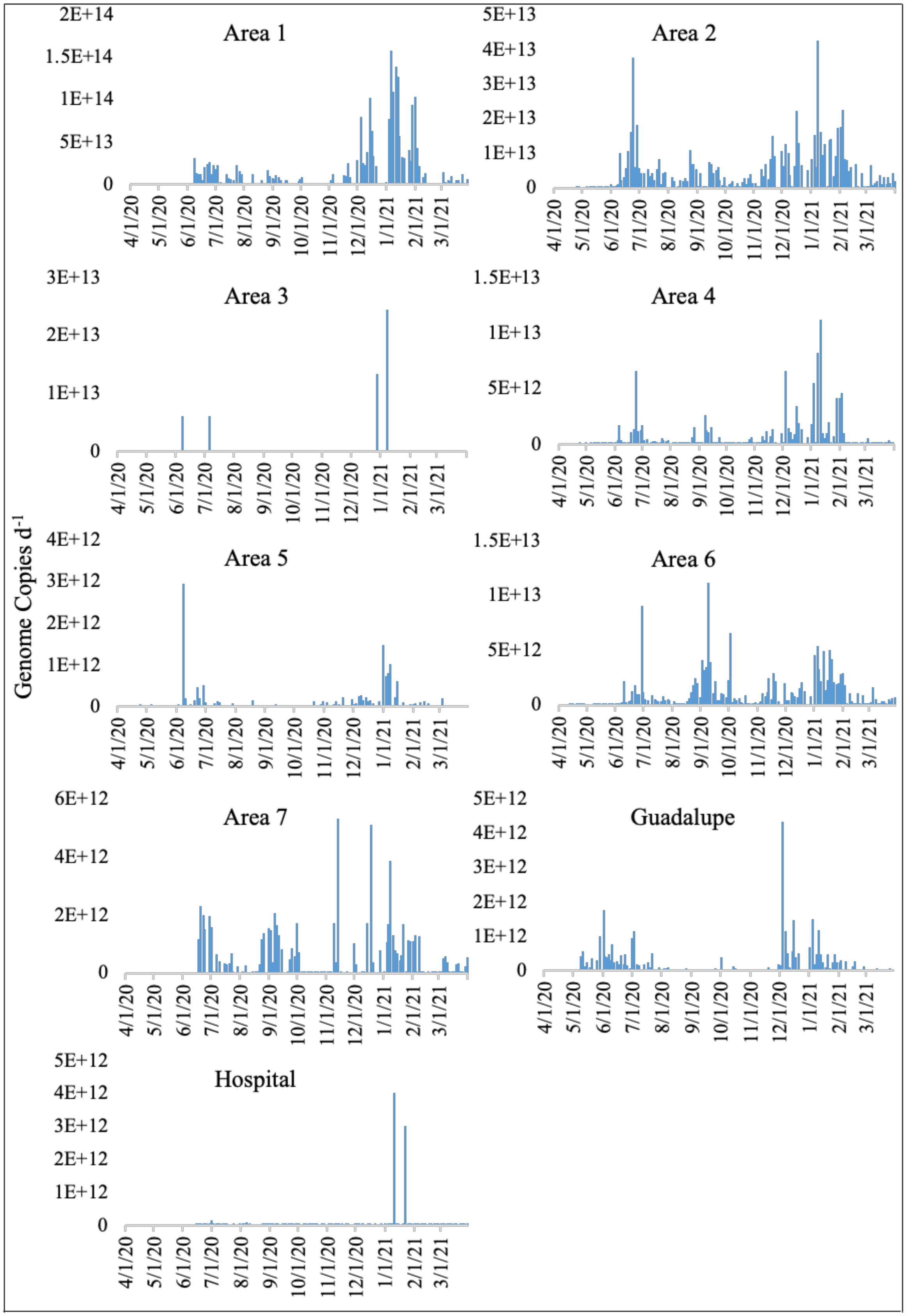


**Figure S4**. Viral load per day (genome copies $d^{-1}$) in target catchments from 1 April 2020 - 31 March 2021.

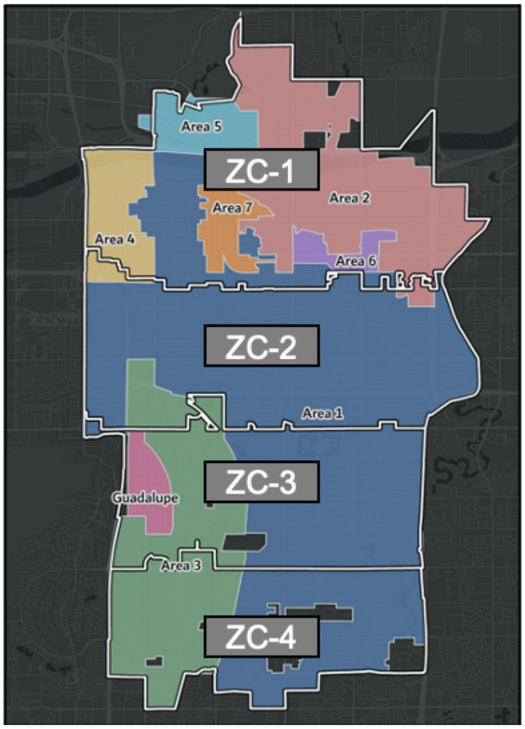


**Figure S5**. Wastewater catchment areas overlapped with Tempe, AZ zip codes 85281, 85282, 85283, and 85284 correspond to ZC-1, ZC-2, ZC-3, ZC-4 informing percent contribution.
