## Supplementary material for "Unrestricted Online Sharing of High-frequency, High-resolution Data on SARS-CoV-2 in Wastewater to Inform the COVID-19 Public Health Response in Greater Tempe, Arizona": Methods

### *Study Location*

This study was conducted within the City of Tempe, Arizona and the Town of Guadalupe, Arizona, (i.e., Greater Tempe), with an estimated residential population of approximately 200,000, and home to Arizona State University, one of the largest public universities in the US. The City was divided into nine sewer catchment areas predetermined by the City for regulatory compliance monitoring purposes and for ease of access in the scale-up of this project. Two additional sampling locations were also included to isolate Tempe-only SARS-CoV-2 signals.

### *Sample Collection*

Flow- and time-weighted 24-h composite samples of untreated wastewater were collected at each sampling location within the wastewater collection system for three days each week (Tuesday, Thursday, Saturday), beginning April 2020 (Catchment 7 and Tempe St. Luke's Hospital were added in July 2020). Samples were collected either with an Avalanche refrigerated sampler or a portable sampler (Teledyne ISCO, Lincoln, NE) using a mixture of wet and dry ice for cooling. Units were equipped with 9 mm inner diameter (ID) silicon tubing on the pump head, and silicon PTFE lined tubing of the same diameter. Flow was monitored by an ISCO LaserFlow meter (Teledyne ISCO, Lincoln, NE), located within a nearby manhole or flow estimated based on historic data. Composite samples were collected in acid-washed bottles and transferred to high-density polyethylene bottles that were quickly placed on ice in coolers for transport. Samples were processed immediately same-day to minimize degradation losses.

### *Sample processing and analysis*

Raw wastewater samples were analyzed for SARS-CoV-2 RNA following sequential steps of filtration, concentration, nucleic acid extraction, and reverse transcriptase quantitative polymerase chain reaction (RT-qPCR) analysis. Approximately 150 mL aliquots of raw wastewater samples were filtered through a sterile 0.45 µm polyether sulfone (PES) membrane filter unit (Fisher Scientific, Lenexa KS) by vacuum for removal of large debris. The filtrate was then loaded onto two Amicon® Ultra 15 centrifugal filters (Millipore Sigma, Burlington, MA) with a 10,000 molecular weight cutoff and centrifuged at ~4,000 RPM for 15 to 20 minutes for five sequential intervals with an Eppendorf 5810R swing bucket refrigerated centrifuge (Eppendorf, Enfield, CT). The final concentrate was combined into 1.5 mL conical microcentrifuge tubes, and 200 µL was processed using an RNeasy mini extraction kit from Qiagen (Germantown, MD), modified for use with this specific matrix following an animal cells protocol. The extracted RNA (50 µL) was stored at -80°C until quantification by RT-qPCR using SuperScript III Platinum One-Step qRT-PCR Kit (Invitrogen, Carlsbad, CA). The Charité/Berlin (World Health Organization) designed primers and probe for the E (envelope) SARS-CoV-2 gene target were purchased from Integrated DNA Technologies (Coralville, IA) [1, 2]. For quality assurance and quality control, deionized water was used for whole process negative extraction controls for each sample batch and RNAase/DNAase free Ultrapure™ water (Invitrogen,Waltham, MA) was used as molecular negative template control along with a SARS-CoV-2 positive control in every RT-qPCR plate. The positive control was created by *in vitro* transcription using linearized plasmids, with subsequent sequencing to determine validity. Later this transitioned to a commercially available synthetic full genome target provided by Twist Bioscience (San Francisco, CA). Standard curves ranged from $10^0$ to $10^6$, with a detection limit cutoff for quantification based on the standard curve (~100 copies $uL^{-1}$). Triplicate standard curves were analyzed for each new batch of assay reagents and were used to quantify samples. Quantification was performed using an Applied Biosystems QuantStudio™ 3 Real-Time

PCR System with the QuantStudio Design and Analysis Software 1.2 from Thermo Scientific (Waltham, MA). Method details are previously published [3].

*Population Estimates*

Resident populations for each sewershed were estimated using 2010 census block group data. Employment estimates were obtained from the Maricopa Association of Governments (MAG) 2019 employment data and included the following classifications: employees living outside of Tempe (non-resident, employed) and Tempe residents (resident, employed). To correct for changes in employment numbers during lockdown events (commercial closures) and telecommuting activities, we used available MAG average weekday traffic volume (compared to normal conditions) in Maricopa County. This percentage was used to correct the non-resident (Tempe employed) employment numbers. Student population estimates were obtained from publicly-available campus resident data and estimates using changes in wastewater flow volume.

*Clinical Data*

Newly detected clinical cases by zip code within the City of Tempe, Arizona were reported daily by the Arizona Department of Health Services. The City of Tempe began extracting and archiving these data on 23 May 2020. Prior to this, daily case data are not available (data are in aggregate as total cases from the start of the pandemic). Maricopa County-level new positive cases, COVID-related hospitalizations, deaths and long-term care facility deaths per day are publicly available and were collected from the Maricopa County Epidemic Curve Dashboard [4].

*Data and Statistical Analysis*

Measured concentrations in each sewer catchment were transformed to viral load (VL) per day (genome copies $d^{-1}$) using the following equation:

$$Viral\ load\ (genome\ copies\ d^{-1}) = C_x\ x\ Q_x \qquad [1]$$

where $C_x$ (genome copies $L^{-1}$) is the measured concentration in a given catchment, $Q_x$ is the total daily volumetric flow rate (L $d^{-1}$). In cases where one sewer catchment flowed into another, viral loads were subtracted to isolate the individual catchments.

Statistical assessments were conducted in MATLAB R2021a (MathWorks, Natick, MA). Root mean square error (RMSE) was used to calculate the offset between different compared data categories using the following equation:

$$RMSE = \sqrt{\sum_{i=1}^{n}(x_i - y_i)}^2 \quad [3]$$

where $n$ is the number of observations, $x_i$ the viral loadings of SARS-CoV-2 in wastewater, and $y_i$ either the newly detected clinical cases, COVID-related hospitalizations, or COVID-related deaths. Data were assessed between 1 April 2020 and 31 March 2021, using individual waves of infection corresponding to up to three events peaking in June/July 2020, August 2020, December/January 2020-21. Data were shifted from 0 to 20 days in both directions for each of the comparisons. The data resolution between clinical cases and wastewater testing were different (daily vs. 3x per week), so clinical results that did not have a corresponding wastewater data point were omitted from the assessment, post shift.
